# Spatiotemporal Transmission of Influenza in the US during the 2022/23 Season

**DOI:** 10.1101/2025.07.10.25331274

**Authors:** Alexia Couture, Matthew Biggerstaff, Michael Sheppard, Alicia Budd, Aaron Kite-Powell, Sinead E. Morris

## Abstract

Understanding the spatiotemporal dynamics of seasonal influenza spread across the United States (US) is crucial for informed public health planning. We explored patterns of influenza transmission during the 2022/23 season in the US and used a mathematical model to infer potential drivers and underlying mechanisms. Leveraging emergency department visit data, we first estimated the timing of influenza onset for the 2022/23 season at the Health Service Area (HSA) level. We then combined the estimated onset times in a gravity-based mechanistic model with covariates that could be associated with influenza spread, including demographics, climate, mobility, and school opening information. We compared multiple models to find the best fit to the onset times, infer factors driving transmission, and identify potential geographic hubs that were most influential in generating early chains of transmission. From the estimated onset times, we found a spatiotemporal pattern that was characterized by early transmission in southern and southeastern HSAs, followed by localized spread to other regions. The best-fit model included absolute humidity and local transmission modulated by school opening times, and four out five potential hubs were located in the southern US (two in Georgia and one each in North Carolina, Texas, and Washington). In conclusion, we found a regional pattern for the spatiotemporal spread of 2022/23 seasonal influenza and identified potential key drivers of this pattern. These findings, and similar studies from other influenza seasons, may improve our understanding of the spatiotemporal spread of influenza in the US.

- We modeled spatiotemporal transmission of influenza virus in the US from 2022–2023.
- Early transmission in the South was followed by localized spread to other regions.
- Population size, absolute humidity and school opening times modulated this spread.
- Hubs in the South and Northwest may have sparked outbreaks in other places.

## Introduction

Seasonal influenza causes substantial morbidity and mortality in the United States (US), with between 9– 40 million symptomatic illnesses, 120,000–700,000 hospitalizations, and 6,000–51,000 deaths estimated to occur each year.^1^ Nationally, influenza seasons typically start in October and peak between December and March the following year. However, there is considerable variability in this timing across seasons and across regions within the US.^2, 3^ The 2022/23 season, which followed an extended period of low influenza activity during the COVID-19 pandemic, was particularly unusual in that it began before October in some regions and peaked as early as November.^4^

Understanding the mechanisms driving different spatiotemporal patterns of influenza spread is important for strengthening seasonal awareness, public health preparedness, and response capabilities. Historically, it has been proposed that seasons tend to start in the southern US (although this pattern is not observed every season), with subsequent transmission dominated by localized spread between nearby regions.^5, 6^ Similar patterns were observed during the 2009 H1N1 influenza pandemic.^7, 8^ Although various drivers have been identified to explain these patterns — including absolute humidity, population demographics, human mobility, and school opening times — differences in their relative importance across studies and time periods complicates efforts to extrapolate findings to more recent seasons.^5-7, 9, 10^

Mathematical models of influenza transmission between regions can capture patterns of spatiotemporal spread and help elucidate the mechanisms underlying observed dynamics. In particular, gravity-based frameworks are designed to capture the increased likelihood of movement (and therefore disease transmission) between more populated regions, and between regions that are closer together.^6, 11, 12^ Such models have been combined with regional estimates of influenza activity onset in the US to determine factors driving spatial spread of the 2002/03 to 2008/09 influenza seasons and the 2009 H1N1 pandemic.^5, 7^ These methods were also extended to identify regions that were most influential in facilitating onward transmission (so-called transmission ‘hubs’) during the 2009 pandemic.^8^ Although the gravity model framework provides a flexible and intuitive method for disentangling the spatiotemporal dynamics of influenza, its effective application requires accurate estimation of the timing of seasonal onset which varies by season.^5^ The 2022/23 influenza season, which was characterized by a distinct and well-defined period of increasing and peak influenza activity, provides an ideal case study for the application of this analytical framework.

In this paper we use highly resolved spatial information on influenza-associated emergency department (ED) visits to explore recent patterns of influenza transmission during the 2022/23 season. We first estimate when the 2022/23 season started across the US, then combine these estimates with a gravity-based framework to infer the factors driving spatial transmission and identify potential hubs. Our findings can help improve our understanding of the patterns and mechanisms underlying the spatiotemporal spread of influenza in the US, particularly in a season with atypical timing.

## Materials and methods

### Data

We used National Syndromic Surveillance Program (NSSP) data to examine ED visits with diagnosed influenza during the 2022/23 influenza season.^13^ NSSP collects electronic health data from approximately 80% of all EDs through a collaboration between CDC, local and state health departments, and healthcare facilities. ED visits are aggregated by Health Service Area (HSA)^14^ and by the Morbidity and Mortality Weekly Report (MMWR)^15^ week in which they occurred, and diagnostic codes are used to identify visits due to influenza. For each HSA, we used the weekly percent of influenza ED visits out of all ED visits as a proxy for influenza activity in time and space. Data were extracted from MMWR week 32 of 2022 to MMWR week 31 of 2023. We note that coverage of NSSP varies across the US, with a considerable area missing from the center of the country that is mostly due to incomplete discharge codes (Figure 1 and S4).

**Figure 1.**
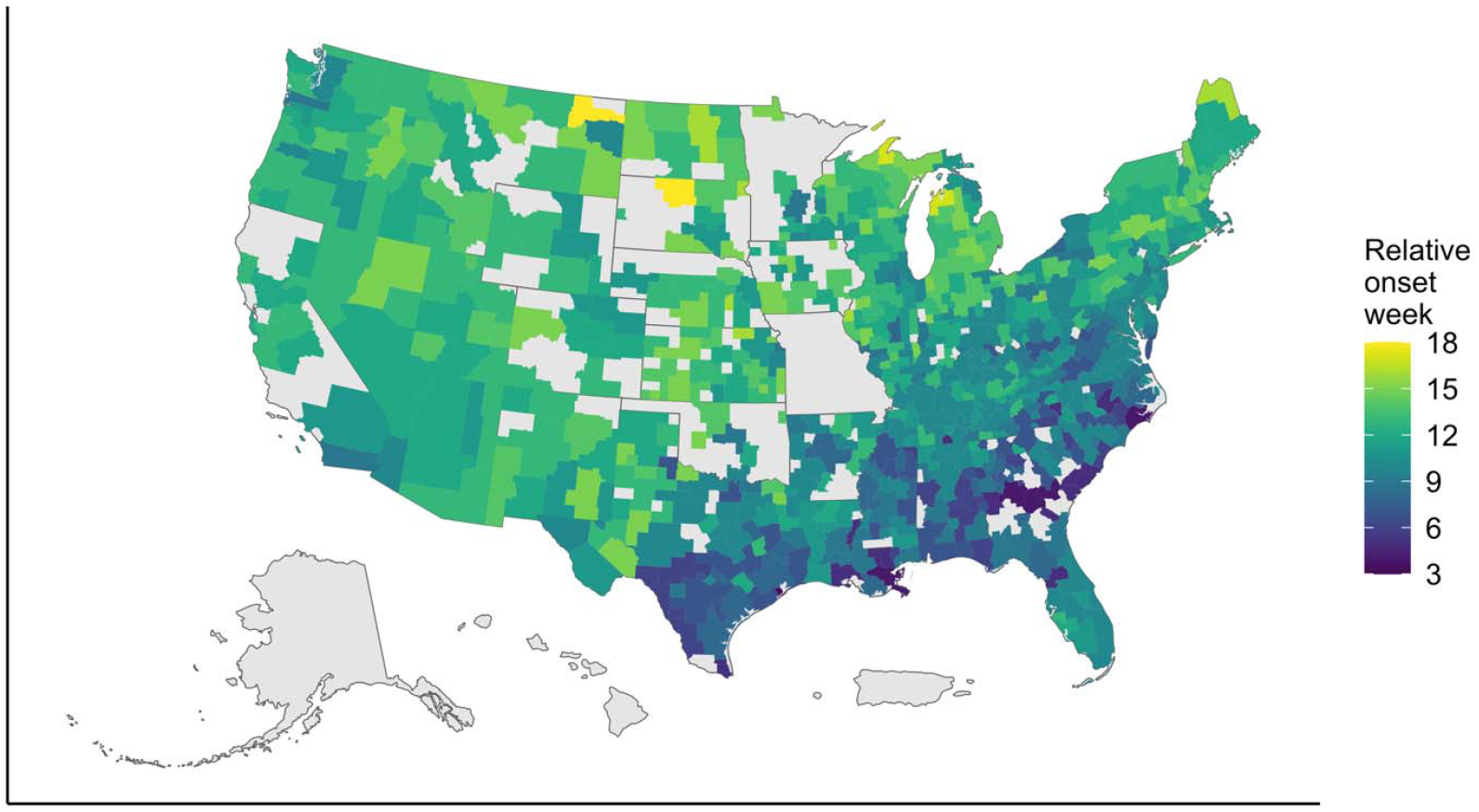
Estimated epidemic onset times for 724 Health Service Areas (HSAs) during the 2022/23 influenza season. Onset weeks are numbered relative to the start of the study period which is MMWR week 32 representing the first week and counting up from then. Regions in grey were either absent from the data or excluded during the analysis.

To reduce the impact of inconsistent NSSP coverage and demographic noise, we excluded HSAs based on the following criteria: 1) those with a population size less than 5,000; 2) those not in the contiguous US; 3) those with an average weekly number of all ED visits per 100,000 population less than 100; 4) those with an average weekly influenza ED visits per 1,000 less than 1; 5) those with less than 3 weeks of nonzero influenza activity before the week of peak activity; 6) those with less than 10 influenza ED visits at the peak; 7) those whose time series failed a Box-Pierce white noise test^16^; and 8) those with a 2 week lag autocorrection less than 0.2. These exclusions removed time series that were without temporal trends or with low to no influenza activity.

We also derived covariates that could be associated with influenza spread from multiple data sources. In brief, these included:

- sociodemographic information on population size, the proportion of the population that is of school age (5–18 years), and the Social Vulnerability Index;^17, 18^
- climatic information on weekly absolute humidity levels;^19^
- mobility data for the weekly number of trips taken in an HSA and the percent of the population staying at home;^20^
- activity of other respiratory viruses monitored by NSSP, defined as the weekly percent of ED visits due to COVID-19 or respiratory syncytial virus (RSV);^13^
- school opening dates from the biggest school district in each state to create a weekly in-session binary variable indicating whether the fall term had started or not; we multiplied this by the proportion of the population that is of school age to scale the effect by HSA;
- spatial data to define the distance (in kilometers) between the centroids of each HSAs^17^.

Data were aggregated weekly and by HSA where possible, and all continuous covariates were scaled. Full details are available in the Supplement.

### Estimating relative onset times

With the NSSP data, we estimated seasonal onset times as the week that influenza activity began to increase in each HSA during the 2022/23 influenza season. We used the breakpoint method, which fits piecewise slopes to each time series from the beginning of the season to the peak, and tests for breakpoints where the slope changes, i.e., when influenza activity starts to increase.^5, 8^ We tested for one or two breakpoints and chose the best fit by minimizing the residual sum of squares.^21^ If two breakpoints had a better fit, then we used the earliest breakpoint to be the estimated onset time. Finally, we undertook an outlier analysis to check for inconsistencies in estimated onset times and re-estimate those that may have occurred too early or too late in relation to the true start of the season (details provided in the Supplement). We also compared the final onset times with the timing of peak influenza activity for each HSA as an alternative metric for epidemic timing and examined the range of time between estimated onset and peak.

Onset times are defined relative to the start of the study period for our main model input, with MMWR week 32 (the week ending August 13, 2022) defined as week 1 for our study. For example, relative onset week 5 (i.e., 5 weeks since the start of our study period) translates to MMWR week 36 (the week ending September 10, 2022). After estimating relative onset times, we conducted descriptive analyses by calculating Spearman correlations between onset times and the covariates described above. We adjusted for multiple testing using the Benjamini-Hochberg correction.

### Spatial transmission model

We investigated the spatial spread of influenza using a gravity-based transmission framework that incorporates terms for local seeding of infection within a HSA and spatial transmission from neighboring HSAs that are already infected.^5-8, 12^ A simplified version of the model is:

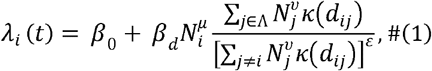

where λ_i_(t) is the force of infection experienced by HSA *i* at time *t,β*_0_ is the local seeding parameter, β_*d*_ is the spatial transmission parameter, *N*_*i*_ is the population size of HSA *i* with corresponding scaling parameter µ, Λ is the set of infected HSAs at time *t, ε* is a normalization term to adjust for population density around HSA i, *N*_*j*_ is the donor population size of HSA *j* with corresponding scaling parameter υ, and *κ* (*d*_*ij*_) is a kernel function describing how the force of infection decays with the distance (*d*_*ij*_)between HSAs *i* and *j*. Overall, the greater the force of infection, the greater the chance an HSA will experience its epidemic onset that week.

We started by fitting various iterations of the model above to the estimated relative onset times.

First, we compared exponential and power-law functions for the distance kernel, defined respectively as:

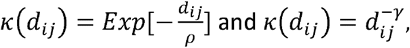

where *ρ* are γ parameters describing how quickly the kernel decays with distance. We also compared the importance of frequency-vs. density-dependent transmission by setting the normalization term ε to 1 or 0, respectively. Finally, we compared the impact of adding donor population size to the gravity-based component of the model by either setting υ to 0 or allowing it to be estimated. We compared model performance using Akaike Information Criteria (AIC) and defined a difference of greater than approximately 2 between models as supporting the model with lower AIC.

Next, we assessed the importance of different covariates by individually including each potential covariate as an additional model component and comparing model performance. We added 1) a climatic component for absolute humidity to the overall force of infection, 2) a school in-session indicator (multiplied by the school age population) in the spatial transmission component alone, the seeding component alone, or in both components, 3) a mobility indicator (the total number of trips taken per population or 1 minus the proportion of the population that stays at home) in the spatial transmission component, the local seeding component, or both components, 4) an SVI indicator in the spatial transmission component, the local seeding component, or both components, and 5) an RSV or COVID-19 indicator in the local seeding component. The expanded model can be expressed as

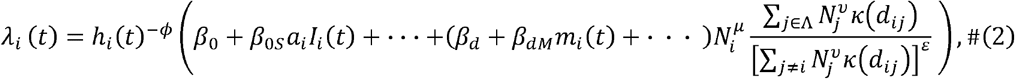

where *h*_*i*_(t) is the absolute humidity in HSA *i* and week t, and *ϕ* is an estimated parameter modulating the relationship between increasing transmissibility and lower absolute humidity that is typically observed in temperate climates.^10, 22^ Different terms can then be added for each new component. For example, the above expression demonstrates how the impact of schools could be incorporated into the local component through the addition of a local seeding transmission parameter when schools are in session (*β*_*0S*_), the school age population for HSA *i* (*a*_*i*_), and the school in-session indicator for HSA *I* and week *t* (*I*_*i*_(*t*)). Similarly, the impact of mobility could be incorporated into the spatial component through the addition of a spatial transmission parameter for mobility (*β*_*dM*_) and the mobility indicator for HSA i and week t (*m*_*i*_ (*t*)).

Model parameters were estimated using maximum likelihood, where the log-likelihood of model parameters Θ given onset times *T*= [*T*_1_, …,*T*_n_] for HSAs1, …,*n* is

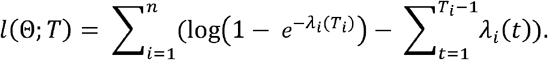

We used Latin-hypercube sampling to generate 20 different initial parameter conditions and then fit each model using the Nelder-Mead simplex algorithm for optimization. Parameter 95% confidence intervals (CIs) were calculated using the Hessian matrix. In addition to comparing AICs, we assessed model fit and parameter identifiability by performing log-likelihood profiling and investigating parameter sensitivity to different initial conditions (further details in the Supplement).

### Identifying Major Influenza Hubs

We followed a previously defined framework to identify potential hubs that were most influential in generating early chains of transmission.^8^ The steps are outlined briefly below and further details can be found in Kissler et al.^8^ First, we note that the mechanistic model can be divided into local seeding and spatial components, where the spatial component combines the force of infection contributed by each HSA at a particular time point. This makes it additive and separable and allows one to derive equations for the force of infection from one HSA to another. For the simplified model (Equation 1), the force of infection from HSA *j* on HSA *i* at ‘s time of onset (*T*_*i*_) is :

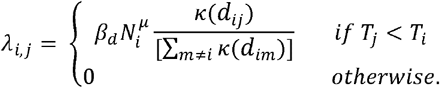

The total force of infection on HSA *i* would then be the sum of the local seeding component and the contribution from all other HSAs:

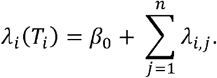

The corresponding probability that location *i* is infected by local seeding (given that it is infected at T_*i*_) 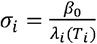 and the probability that it is infected by HSA *j* is 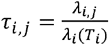.

To reconstruct likely “infection pathways” we first define a reverse transmission matrix from the above probabilities as:

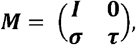

where ***I*** is the identity matrix, **0** is a zero matrix, ***σ*** is a matrix with *σ*_1_, *σ*_2_, ‥, *σn* along the diagonal and zeroes elsewhere, and **τ** is a strictly lower triangular matrix whose-th entry is τ_*i,j*_, where τ_*i,j*_= 0 for *j* ≥ *i*. Since the *p*-th power of ***M*** is the probability of transmission between two HSA’s by *p* − 1 intermediate steps, one can determine the ultimate source of an outbreak for each HSA by finding the limit as *p* tends to infinity, i.e.,

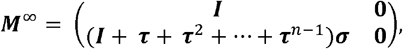

where element (***M* ^∞^**)_*i,j*_ is the probability that state was the ultimate source of the outbre. Hubs are then identified using the lower left block,

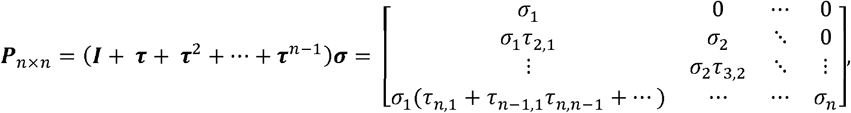

where ***P*** _*i,j*_ is the probability that local seeding in HSA *j* ultimately led to the outbreak in HSA *i* and the column sums of ***P*** can be interpreted as the expected number of outbreaks triggered by each HSA. We applied the above framework to our best-fit model and identified the most likely hubs as HSAs that had the highest probabilities of local seeding and likely triggered infection in the largest number of other HSAs.

All analyses were performed in R version 4.4.0.

## Results

### Patterns of spatiotemporal spread

We estimated onset times for 724 of the 949 HSAs in the US during the 2022/23 influenza season (Figure 1; Figure S3). The HSA for Galveston, TX experienced the earliest onset in MMWR week 34 (i.e., relative onset week 3 with week ending on August 27, 2022), whereas the HSA for Roosevelt – Valley, MT and the HSA for Dewey – Walworth, SD experienced the latest onsets in MMWR week 49 (i.e., relative onset week 18 with week ending December 10, 2022). Across the US, we found a spatiotemporal pattern characterized by earlier onsets in southern and southeastern HSAs, followed by generally localized spread to other regions over the course of 15 weeks, and later onsets typically in northern central and eastern HSAs. The spatiotemporal pattern of peak timing was consistent with this finding (Figure S4). The time between estimated onset week and peak week was heterogeneous across the US, ranging from 1 to 12 weeks (Figure S5).

Across sociodemographic covariates, earlier onset times were associated with larger population sizes (correlation -0.29, p < 0.001; Figure 2A), higher SVI (i.e., greater social vulnerability; correlation - 0.35, p < 0.001; Figure 2B), and earlier school opening times (correlation = 0.17, p < 0.001; Figure 2C). We did not detect an association with the proportion of the population 5–18 years (correlation -0.04, p = 0.32; Figure 2D). When testing mobility metrics, we found that earlier onsets were associated with a greater average number of trips taken in the week prior to onset (correlation -0.29, p < 0.001; Figure 2E) but not with the average proportion of the population staying at home (correlation = -0.01, p = 0.70; Figure 2F). Finally, we found that earlier onset times were associated with greater absolute humidity levels (correlation = -0.61, p < 0.001; Figure 2G) and RSV activity (correlation = 0.31, p < 0.001; Figure 2H) in the week prior to onset, but not COVID-19 activity (correlation = 0.07, p = 0.08; Figure 2I). These latter associations must be interpreted with caution given that (i) absolute humidity is also correlated with geographic location and tends to be higher in the southern US; and (ii) the change in respiratory virus activity in the weeks prior to onset (i.e., whether RSV activity is increasing or declining) may be more informative than a static value from the week prior. More generally, there are limited inferences that can be made from these descriptive analyses due to the potential for interactions between covariates. We therefore sought to systematically test the importance of each covariate by fitting a series of multivariate, gravity-based mechanistic models to the estimated onset times.

**Figure 2.**
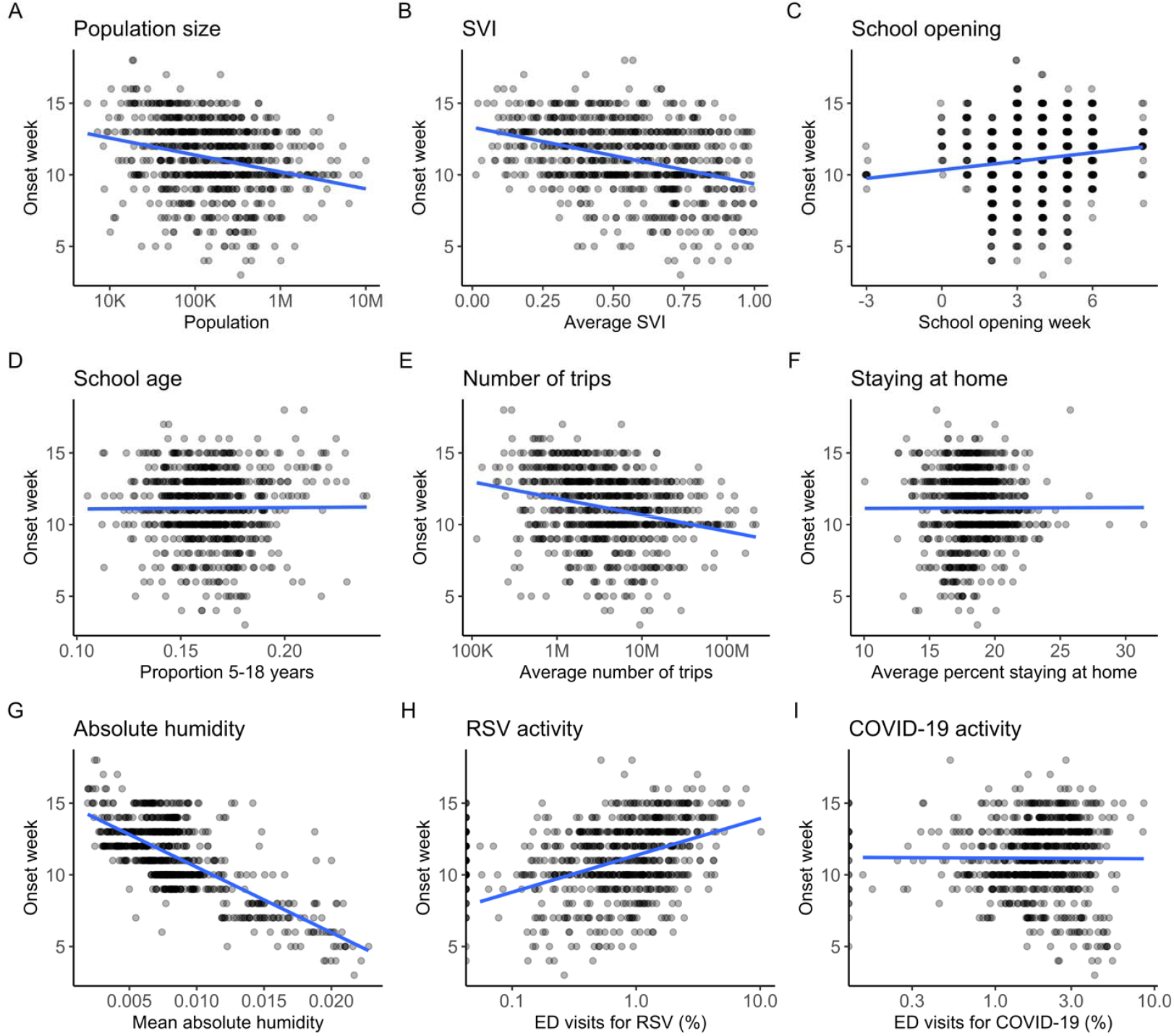
Descriptive associations between select covariates and estimated relative onset times. Trend lines are provided as a visual guide only. Relative onset week 1 is aligned with MMWR week 32 which in ends August 13, 2022.

### Mechanistic drivers of transmission

When fitting different iterations of the simplified model (Equation 1), we found that an exponential distance kernel outperformed a power law distance kernel and frequency-dependent transmission (*ε* = 1) outperformed density-dependent transmission (*ε* = 0) (Table S1). In addition, adding donor population size to the gravity-based component of the model did not improve the fit. Therefore, in all further iterations of the model, we excluded donor population size and assumed frequency-dependent transmission with an exponential distance kernel.

Next, we compared the impact of including each remaining covariate in turn to either the local seeding or spatial transmission components of the model (i.e., the terms), with the exception of absolute humidity which was included through a multiplicative power law (Equation 2). The inclusion of absolute humidity provided a substantially better fit that the inclusion of any other single covariate (Table S1). We therefore fixed absolute humidity in the model and repeated the process above and found that adding school opening times to the local component provided the next greatest improvement in model fit. Adding subsequent covariates did not substantially improve model fit and introduced potential issues with parameter identifiability. We therefore concluded that a model with absolute humidity and local transmission modulated by school opening times provided the most parsimonious explanation of the spatiotemporal distribution of onset times. Parameter estimates for this model are provided in Table 1 and profile likelihoods are shown in Figure S6.

**Table 1.** Fitted parameter estimates (and 95% confidence intervals) from the most parsimonious model.

| Parameter | Associated term or covariate | Estimate (95% confidence interval) |
| --- | --- | --- |
| $\beta_0$ | Baseline local seeding | 0.00035 (0.00004, 0.00272) |
| $\beta_d$ | Baseline spatial transmission | 0.02608 (0.01101, 0.06174) |
| $\mu$ | Host population size | 0.29040 (0.23312, 0.36176) |
| $\rho$ | Exponential distance kernel | 150.116 (129.444, 174.089) |
| $\phi$ | Absolute humidity | 0.35106 (0.21948, 0.56154) |
| $\beta_{os}$ | School age population x school opening indicator | 0.01681 (0.00702, 0.04021) |

### Hubs of infection

Using the most parsimonious model, we identified the ten most likely hubs as the HSAs with the greatest seeding probabilities (Table 2). The hub with the greatest seeding probability and expected number of outbreaks was Galveston, TX, the HSA with the earliest estimated onset week. Of the five next most likely hubs with seeding probabilities ≥ 99%, four were located in the Southeast (one in North Carolina and three in Georgia) and one in the Northwest (in Washington). These hubs were not necessarily HSAs with the largest population sizes or earliest onset times.

**Table 2.** Ten most likely hubs identified using the most parsimonious model. Hubs are ordered by seeding probability (which was subsequently rounded to two decimal places) following by expected number of outbreaks.

| HSA | Seeding probability | Expected outbreaks | MMWR onset week | Relative onset week | Population |
| --- | --- | --- | --- | --- | --- |
| Galveston, TX | 1.000 | 131 | 34 | 3 | 342,139 |
| Onslow – Craven, NC | 1.000 | 88 | 35 | 4 | 391,695 |
| Lewis – Pacific, WA | 1.000 | 43 | 40 | 9 | 103,178 |
| Bulloch – Emanuel, GA | 0.997 | 84 | 35 | 4 | 127,023 |
| Laurens – Toombs, GA | 0.995 | 77 | 35 | 4 | 144,412 |
| Bibb (Macon), GA - Houston, GA | 0.992 | 90 | 35 | 4 | 456,389 |
| San Diego (San Diego) – Imperial, CA | 0.881 | 19 | 40 | 9 | 3,519,545 |
| Hennepin (Minneapolis) - Anoka, MN | 0.522 | 14 | 40 | 9 | 2,192,126 |
| Lincoln, TN | 0.493 | 24 | 36 | 5 | 34,366 |
| Iredell (Statesville) - Alexander, NC | 0.269 | 17 | 36 | 5 | 219,303 |

Finally, we explored the potential spatial influence of the five hubs with greatest seeding probabilities (Figure 3). There was distinct spatial partitioning in the influence of each hub, with outbreaks in the south and central US most likely the result of transmission chains originating from Galveston, TX, and outbreaks in the Northeast most likely the result of transmission stemming from the hubs in North Carolina or Georgia. We also detected a smaller region in the Northwest with transmission likely originating from Lewis – Pacific, WA. These findings highlight the importance of localized spatial transmission in determining patterns of influenza spread during the 2022/23 season.

**Figure 3.**
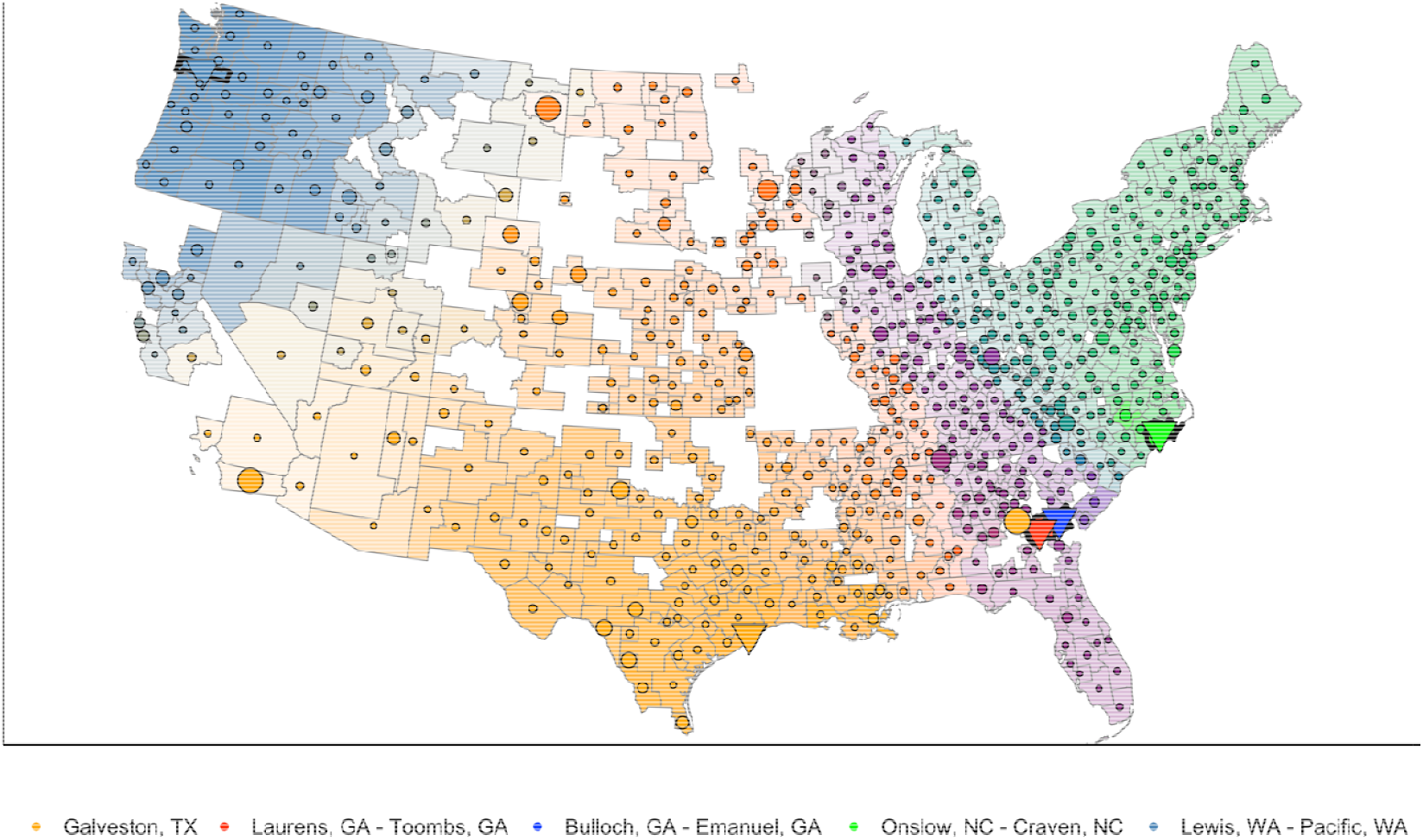
Potential influence of five most likely hubs with the greatest seeding probability. Triangles show hub locations. For all other Health Service Areas (HSAs), larger circle size represents greater seeding probability, and color shade and intensity depicts the likelihood that a particular hub was the ultimate source of infection for that HSA, relative to the next most likely source hub. For color shade, top two hubs were mixed based on proportion of influence on each HSA.

## Discussion

The 2022/23 US influenza season began atypically early and marked the return to significant and sustained influenza transmission since the COVID-19 pandemic (following low levels of transmission in 2020/21 and 2021/22).^3^ By modeling spatially resolved data on ED visits for influenza, we found that the season was characterized by localized spatial spread that originated in southern and southeastern HSAs and was mediated by host population size, absolute humidity, and school opening times. We also identified potential hubs in Texas, Georgia, North Carolina, and Washington that may have been the ultimate source of outbreaks in many other HSAs. Together, these results update and enhance our understanding of the patterns and mechanisms underlying the spatiotemporal spread of influenza in the US.

Our finding that outbreaks originated in the South, subsequently spread in a localized manner, and ended in northern central and northeastern regions echoes patterns observed during the 2009 H1N1 pandemic and a number of influenza seasons prior to 2009.^5, 7^ Charu et al. found seasonal influenza activity often starts in southern states and follows a localized spatial trajectory, and Gog et al. found a similar trend during the 2009 H1N1 pandemic with spread initiating in southern states and subsequently moving north and west. This suggests that recent patterns of influenza spread are still consistent with those identified over a decade ago. Although spatiotemporal spread following the 2009 H1N1 pandemic is less-well characterized at fine spatial resolutions, state-level analysis of the 2010/11– 2019/20 influenza seasons has highlighted earlier peak timing among southern and southeastern states, again consistent with our findings for 2022/23.^5, 10^ We note that the distinctive progression of the 2022/23 influenza season, with its well-defined peak of activity across the US, provided a unique opportunity to evaluate spatial spread with a gravity-based model and offered insights on the spatiotemporal transmission of influenza that may not be as identifiable in other seasons. For example, estimating onset times for seasons with dual peaks or less well-defined increases to peak activity would be more complex and introduce greater uncertainty in subsequent model inferences.

In terms of drivers of transmission, we identified important roles for absolute humidity, which has been associated with the onset of seasonal epidemics in the US^5, 10^, and host population size and school opening times, which were identified as key factors driving the spatiotemporal spread of the 2009 H1N1 pandemic.^7, 8, 23^ Although school opening times have not demonstrated historically strong associations with seasonal influenza transmission, it is possible that the relationship identified here was facilitated by the earlier onset of the 2022/23 season, which was more aligned with the start of school term compared to typical influenza seasons.^5, 10^ However, information on school term dates at finer spatial resolution would be required to investigate this more fully.

In addition to establishing key drivers of influenza transmission, we also identified potential hubs that may have been most influential in facilitating spatial spread across the country. As with prior work^8^, these hubs were not exclusively HSAs with large population sizes or with the earliest estimated onset times. In particular, one potential hub in the Northwest, the Lewis – Pacific HSA in Washington, had an estimated onset 6 weeks after the first estimated onset of the Galveston HSA in Texas. This could reflect an independent seeding event, either following external introduction from outside the US or long-range domestic transmission, that facilitated subsequent spread in the western US. The precise location of hubs were different to those identified during the 2009 H1N1 pandemic, likely due in part to different spatial aggregations used for analysis and inherent inter-season variability in influenza transmission. However, there were general similarities at the state and regional level, with both analyses highlighting hubs with high seeding probabilities in the South (e.g., Georgia), and West (e.g., California). As previous analyses for seasonal influenza have not characterized potential hubs, future work exploring trends over longer timescales could provide important insights into whether such similarities remain consistent from season to season.

There are a number of limitations to our analysis. First, we were not able to estimate onset times for all HSAs in the US and had particularly low coverage in California and the central US. This could have influenced our estimated spatial kernel by requiring longer-range transmission to connect HSAs bordering regions with missing information. We also cannot rule out the possibility that there were influential hubs among the HSAs not included in our analysis. Second, the breakpoint method for estimating onset times is subject to error, particularly when applied to data at fine spatial resolution with greater amounts of noise. However, we developed a number of criteria to exclude noisy time-series and refine suspect onset estimates and found that this method still performed better than other commonly used methods for estimating epidemic onsets. Relatedly, we estimated onset weeks using ED visit data which do not capture all influenza cases in an HSA. Although this may impact the magnitude of observed epidemics, it should not impact estimates of epidemic onset unless the proportion of cases diagnosed through ED visits changed substantially during the weeks surrounding the onsets. Third, we note a greater likelihood of encountering parameter identifiability issues when fitting models of increasing complexity. Although our best-fitting model represents the most parsimonious explanation of the available data and identifies key factors influencing influenza transmission, it is possible that there are additional factors with more subtle influences that we did not identify due to increased model complexity. Fourth, we used population size as our primary demographic covariate. Alternative metrics, such as population density, may better incorporate the variability in HSA size and how populations are distributed within HSAs across the US. However, our finding that frequency-dependent transmission was favored over density-dependent transmission suggests that the inclusion of such metrics would not improve the best-fit mechanistic model. Finally, it is important to highlight that 2022/23 was an atypically early season that occurred following an extended period of low influenza transmission during the first two years of the COVID-19 pandemic. As such, there may have been other factors modulating influenza transmission that were unique to this season, including lower levels of population immunity and changes in individual behavior, that mean our results may not necessarily be generalizable to other seasons.

In this paper we modeled spatiotemporal patterns of influenza transmission during the 2022/23 season using fine-scale spatial information on US influenza-associated ED visits. Overall, we found patterns of spread and important drivers of influenza transmission that were generally consistent with those identified during prior seasons and the 2009 H1N1 pandemic. Our findings provide an updated picture of the spatiotemporal spread of seasonal influenza in the US and can enhance our understanding of the factors that are most influential in driving transmission.

## Data Availability

Data on influenza-associated emergency department visits may be available from the authors upon reasonable request. All other data used in this study are publicly available and their sources are cited in the text.

## Acknowledgements

State, local, and territorial health departments participating in CDC’s National Syndromic Surveillance Program.

## References

1. CDC. Flu Disease Burden: Past Seasons 2025 [Available from: https://www.cdc.gov/flu-burden/php/data-vis/past-seasons.html.

2. CDC. Flu Season 2024 [Available from: https://www.cdc.gov/flu/about/season.html.

3. CDC. FluView Interactive 2025 [Available from: https://www.cdc.gov/fluview/overview/fluview-interactive.html.

4. CDC. Influenza Activity in the United States during the 2022-2023 Season and Composition of the 2023-2024 Influenza Vaccine 2023 [Available from: https://www.cdc.gov/flu/whats-new/22-23-summary-technical-report.html.

5. Charu V, Zeger S, Gog J, Bjornstad ON, Kissler S, Simonsen L, et al. Human mobility and the spatial transmission of influenza in the United States. PLoS Comput Biol. 2017;13(2):e1005382.

6. Viboud C, Bjornstad ON, Smith DL, Simonsen L, Miller MA, Grenfell BT. Synchrony, waves, and spatial hierarchies in the spread of influenza. Science. 2006;312(5772):447–51.

7. Gog JR, Ballesteros S, Viboud C, Simonsen L, Bjornstad ON, Shaman J, et al. Spatial Transmission of 2009 Pandemic Influenza in the US. PLoS Comput Biol. 2014;10(6):e1003635.

8. Kissler SM, Gog JR, Viboud C, Charu V, Bjornstad ON, Simonsen L, et al. Geographic transmission hubs of the 2009 influenza pandemic in the United States. Epidemics. 2019;26:86–94.

9. Dalziel BD, Kissler S, Gog JR, Viboud C, Bjornstad ON, Metcalf CJE, et al. Urbanization and humidity shape the intensity of influenza epidemics in U.S. cities. Science. 2018;362(6410):75–9.

10. Shaman J, Pitzer VE, Viboud C, Grenfell BT, Lipsitch M. Absolute humidity and the seasonal onset of influenza in the continental United States. PLoS Biol. 2010;8(2):e1000316.

11. Xia Y, Bjornstad ON, Grenfell BT. Measles metapopulation dynamics: a gravity model for epidemiological coupling and dynamics. Am Nat. 2004;164(2):267–81.

12. Eggo RM, Cauchemez S, Ferguson NM. Spatial dynamics of the 1918 influenza pandemic in England, Wales and the United States. J R Soc Interface. 2011;8(55):233–43.

13. CDC. National Syndromic Surveillance Program (NSSP): About NSSP 2024 [Available from: https://www.cdc.gov/nssp/php/about/index.html.

14. Makuc DM, Haglund B, Ingram DD, Kleinman JC, Feldman JJ. Health service areas for the United States. Vital Health Stat 2. 1991(112):1–102.

15. CDC. MMWR Weeks 2025. Available from: https://ndc.services.cdc.gov/wp-content/uploads/MMWR_week_overview.pdf.

16. Box GEP, Pierce DA. Distribution of Residual Autocorrelations in Autoregressive-Integrated Moving Average Time Series Models. Journal of the American Statistical Association. 1970;65(332):1509–26.

17. tidycensus: Load US Census Boundary and Attribute Data as ‘tidyverse’ and ‘sf’-Ready Data Frames [Internet]. 2025. Available from: https://walker-data.com/tidycensus/.

18. Place and Health - Geospatial Research, Analysis, and Service Programs (GRASP): SVI Data & Documentation Download [Internet]. 2020. Available from: https://www.atsdr.cdc.gov/place-health/php/svi/svi-data-documentation-download.html.

19. Datasets [Internet]. 2025 [cited 14 May 2024]. Available from: https://cds.climate.copernicus.eu/.

20. Trips by Distance [Internet]. 2024 [cited 14 May 2024]. Available from: https://data.bts.gov/Research-and-Statistics/Trips-by-Distance/w96p-f2qv/about_data.

21. Zeileis A, Leisch F, Hornik K, Kleiber C. strucchange: An R Package for Testing for Structural Change in Linear Regression Models. Journal of Statistical Software. 2002;7(2):1 – 38.

22. Shaman J, Kohn M. Absolute humidity modulates influenza survival, transmission, and seasonality. Proc Natl Acad Sci U S A. 2009;106(9):3243–8.

23. Chao DL, Halloran ME, Longini IM, Jr. School opening dates predict pandemic influenza A(H1N1) outbreaks in the United States. J Infect Dis. 2010;202(6):877–80.

