## Supplemental File for "Spatiotemporal Transmission of Influenza in the US during the 2022/23 Season"

**Supplementary Information**

Covariate data

*Population size and proportion of school age*

We obtained population size estimates from 2019 for each US county using the tidycensus package in R and aggregated these to the HSA level (Figure S1A) [1]. Similarly, we aggregated county-level information from 2020 on the total number of people and total number of children aged 5–18 years to generate estimates of the proportion of each HSA population that was of school age (Figure S1B).

*School opening times*

School data was pulled from the webpage of the biggest school district in each state to create a weekly binary variable of in-session or not to represent the start of school. This was used as a proxy for each HSA and was therefore the same across each state. We multiplied this by proportion of school age to adjust effect by HSA.

(Figure S1C)

*Social Vulnerability Index (SVI)*

County-level SVI percentile rankings from 2022 were obtained from the Centers for Disease Control and Prevention (CDC) / Agency for Toxic Substances and Disease Registry (ATSDR) Geospatial Research, Analysis, and Services Program [2]. We aggregated these to the HSA-level by calculating the population-weighted average across counties within each HSA (Figure S1D).

*Absolute humidity*

Climate data were obtained from the ERA5 reanalysis dataset in the Copernicus Climate Change Service (C3S) Climate Data Store and included 2 meter temperature and dew point temperature data for the continguous US at 6-hourly intervals from 2022–2023 [3]. We calculated absolute humidity from these variables and then derived mean values for each week of the 2022/23 season [4]. The original data were provided on a 0.25 x 0.25° latitude/longitude grid and were matched to HSAs by assigning values from the grid point closest to the centroid of each HAS (Figure S2A–B).

*Mobility*

We obtained county-level mobility data from the US Department of Transportation, Bureau of Transport Statistics [5]. These data are based on anonymized mobile phone records and include daily estimates of the total number of trips taken by the county population (where trips are defined as movements away from home that included a stay of more than 10 minutes), and the number of people who stayed at home (i.e., made no trips more than a mile away from home). We aggregated these data to the HSA level and calculated the mean number of trips and proportion of the population staying at home for each week in our study period (Figure S2C–D).

*RSV and COVID-19 activity*

Similar to the influenza data, information on RSV and COVID-19 activity was collated from the National Syndromic Surveillance Program (NSSP) and consisted of weekly emergency department visits for RSV and COVID-19, in addition to the total number of emergency department visits, reported by participating facilities in each HAS [6]. We calculated the proportion of visits due to RSV and COVID-19 for each HSA and week in our study period (Figure S2E–F).

Identification and re-estimation of outlying epidemic onset times

We undertook an outlier analysis to check for inconsistencies in estimated onset times, i.e., to identify onset estimates that may have occurred too early or too late in relation to the true start of the season. First, we identified HSAs whose time-series may have been impacted by lingering transmission from the 2021/22 season as those with 3 weeks of consecutive decreases in influenza activity at the beginning of 2022/23. We re-estimated onset times after removing those first 3 weeks. We also identified HSAs with an initial onset estimate that was potentially too early or too late as those with 4 weeks of consecutive decreases after the estimated onset or 4 weeks of consecutive increases before the estimated onset, respectively. No HSAs were identified as potentially being too early and visual inspection suggested many of those identified as potentially being too late had accurate onset estimates. We therefore identified a subset of these latter HSAs for which the estimated onset occurred when influenza activity was already relatively high (defined as activity greater than approximately 20% of activity reached at the peak). Many of these HSAs had local peaks in influenza activity occurring before the main peak of the season, and so we re-estimated onset times after truncating each time series to the latest local peak. Onset estimates for HSAs without local peaks that could not be re-estimated were discarded from the analysis.

Likelihood profiling

We conducted likelihood profiling to assess model convergence and parameter identifiability. After fitting each model, one-dimensional profiling was performed by varying each parameter in turn and recalculating the log-likelihood (while keeping all other parameters fixed at their estimated values). Two-dimensional profiling was performed by varying two parameters simultaneously while keeping all other parameters fixed. The latter was used to identify potential correlations between fitted parameters.

Supplementary Tables

**Table S1. Model comparisons.** AIC values (ΔAIC) are quoted relative to the minimum AIC value across all models. The model with ΔAIC = 0 is the model with lowest AIC.

| **Model** | Δ**AIC** |
| --- | --- |
| **Simplified model with:** |  |
| Exponential kernel and *ε* = 1 | 18.95 |
| Power law kernel and *ε* = 1 | 25.57 |
| Exponential kernel and *ε* = 0 | 338.60 |
| Exponential kernel, *ε* = 1, and donor population size | 18.29 |
| **Simplified model with exponential kernel,** *ε* **= 1, and:** |  |
| Absolute humidity | 4.29 |
| School age x school indicator (local) | 14.74 |
| School age x school indicator (spatial) | 20.88 |
| School age x school indicator (both) | 16.77 |
| Social Vulnerability Index (local) | 16.65 |
| Social Vulnerability Index (spatial) | 20.96 |
| Social Vulnerability Index (both) | 18.80 |
| Percent staying at home (local) | 20.69 |
| Percent staying at home (spatial) | 20.96 |
| Percent staying at home (both) | 22.87 |
| Total trips (local) | 20.96 |
| Total trips (spatial) | 18.31 |
| Total trips (both) | 20.48 |
| RSV activity (local) | 18.16 |
| COVID-19 activity (local) | 20.96 |
| **Simplified model with exponential kernel,** *ε* **= 1, absolute humidity, and:** |  |
| School age x school indicator (local) | **0.00** |
| School age x school indicator (spatial) | 6.37 |
| School age x school indicator (both) | 2.36 |
| Social Vulnerability Index (local) | 1.53 |
| Social Vulnerability Index (spatial) | 4.86 |
| Social Vulnerability Index (both) | 3.26 |
| Percent staying at home (local) | 6.62 |
| Percent staying at home (spatial) | 6.68 |
| Percent staying at home (both) | 10.39 |
| Total trips (local) | 6.30 |
| Total trips (spatial) | 4.14 |
| Total trips (both) | 6.64 |
| RSV activity (local) | 2.63 |
| COVID-19 activity (local) | 6.29 |

Supplementary Figures

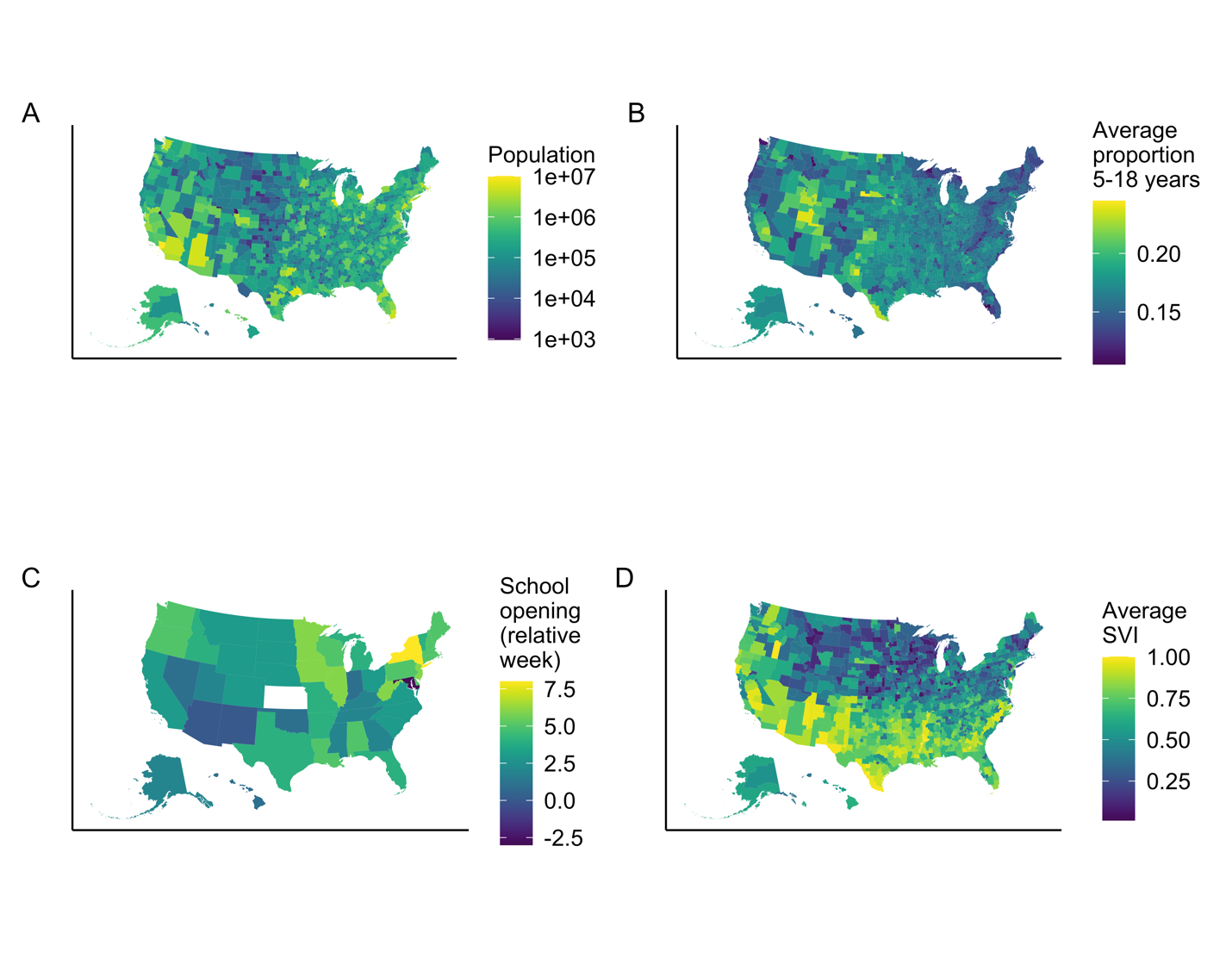
**Figure S1. Sociodemographic covariates.** Shown are covariates for population size (A), the proportion of the population between 5–18 years (B), school opening times relative to the first week of the season (C), and average Social Vulnerability Index (SVI; D).

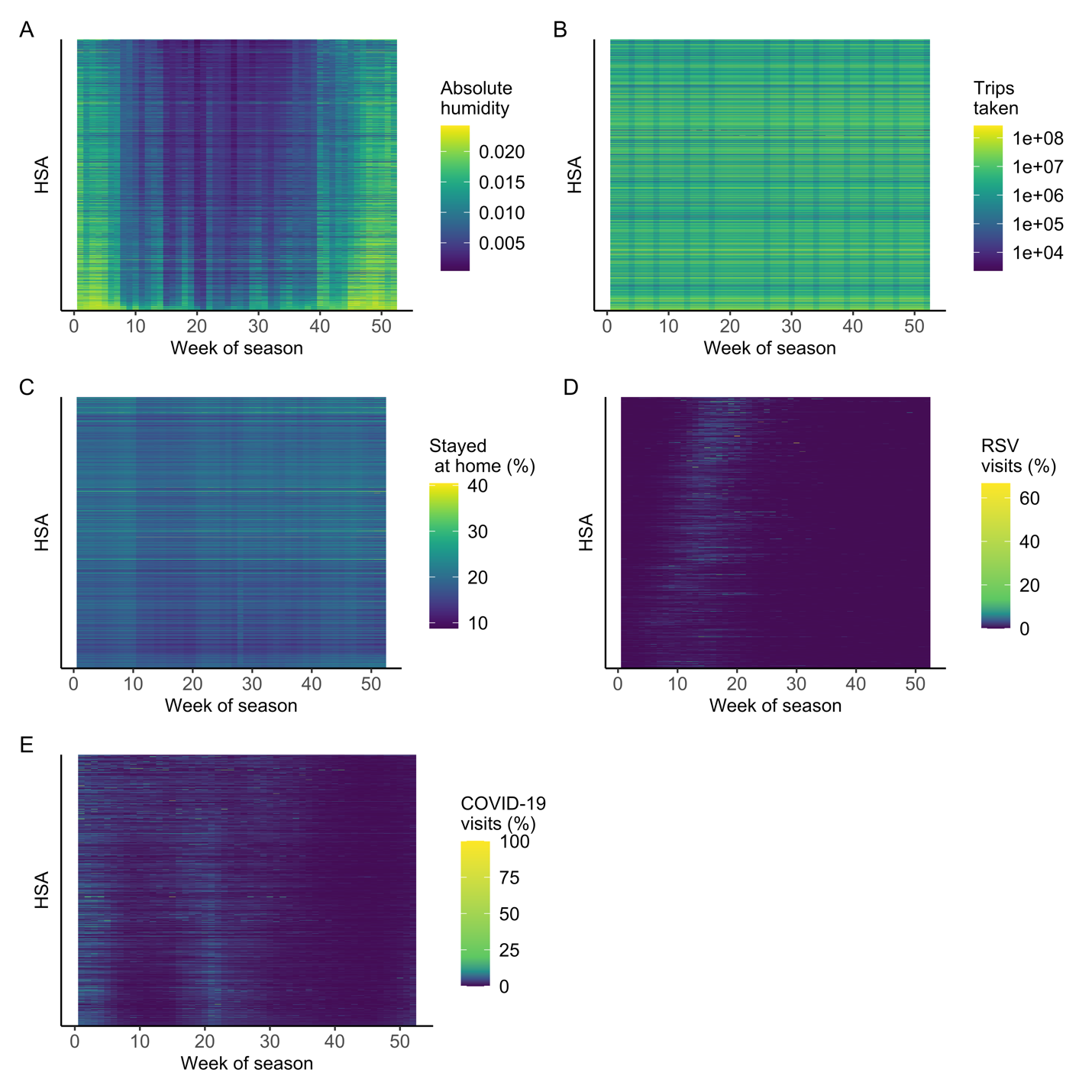
**Figure S2. Time-varying covariates.** Shown are covariates for weekly mean absolute humidity (A), weekly mean temperature (B), weekly mean number of trips taken (C), weekly mean proportion of people staying at home (D), weekly % of emergency department visits for RSV (E), and weekly % of emergency department visits for COVID-19 (F). HSAs are ordered along the y-axis (from north to south) by latitude.

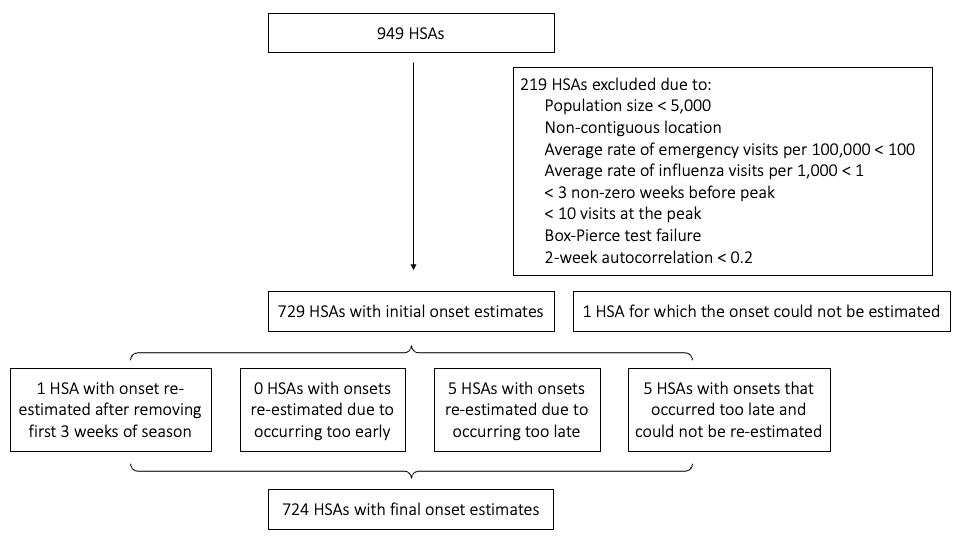
**Figure S3. HSAs included and excluded during estimation of epidemic onset times.**

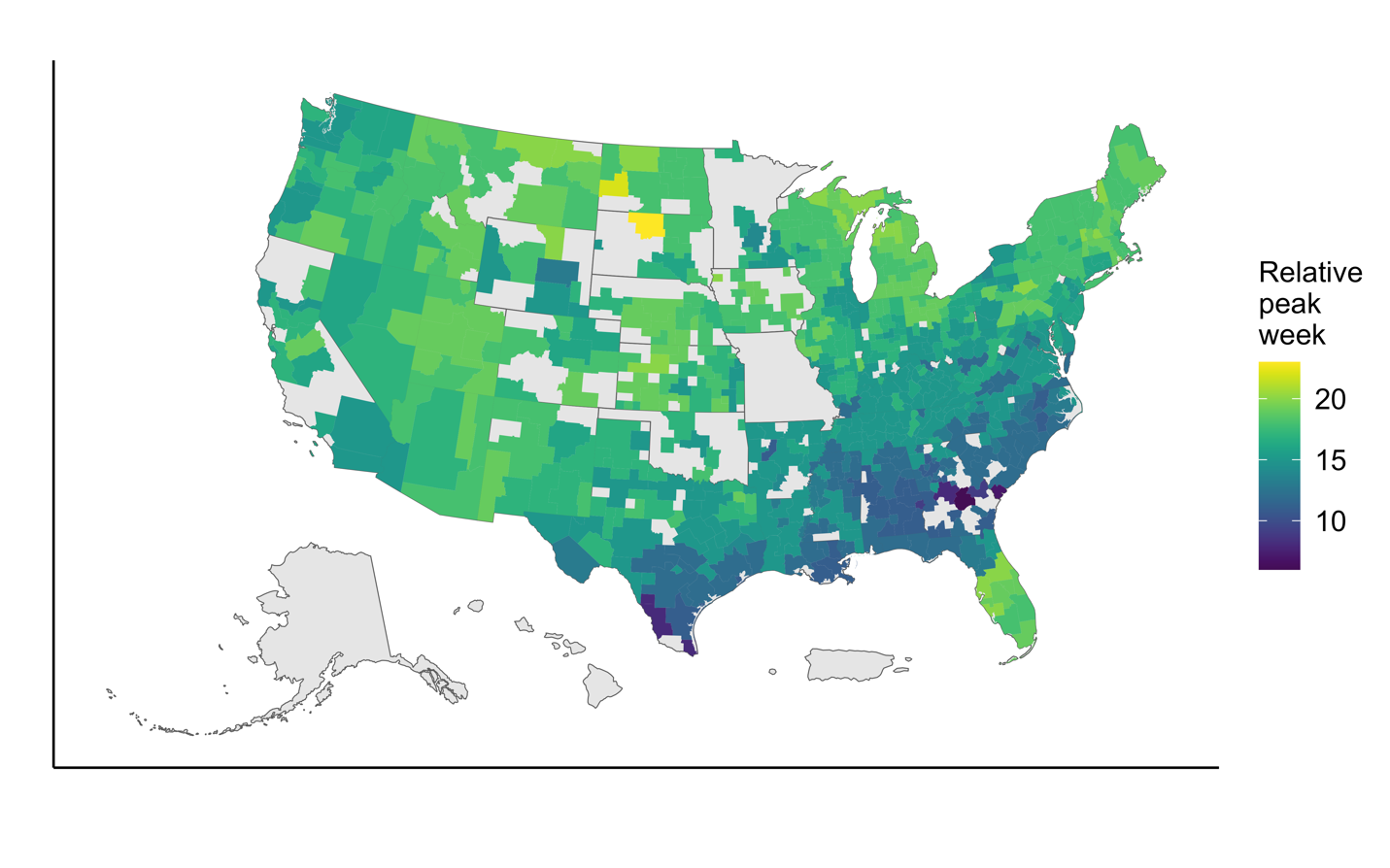
**Figure S4. Estimated peak week of the 2022/23 influenza season for 724 Health Service Areas (HSAs).** Peak weeks are numbered relative to the start of the study period (calendar week 32). Regions in grey were either absent from the data or excluded during the analysis.

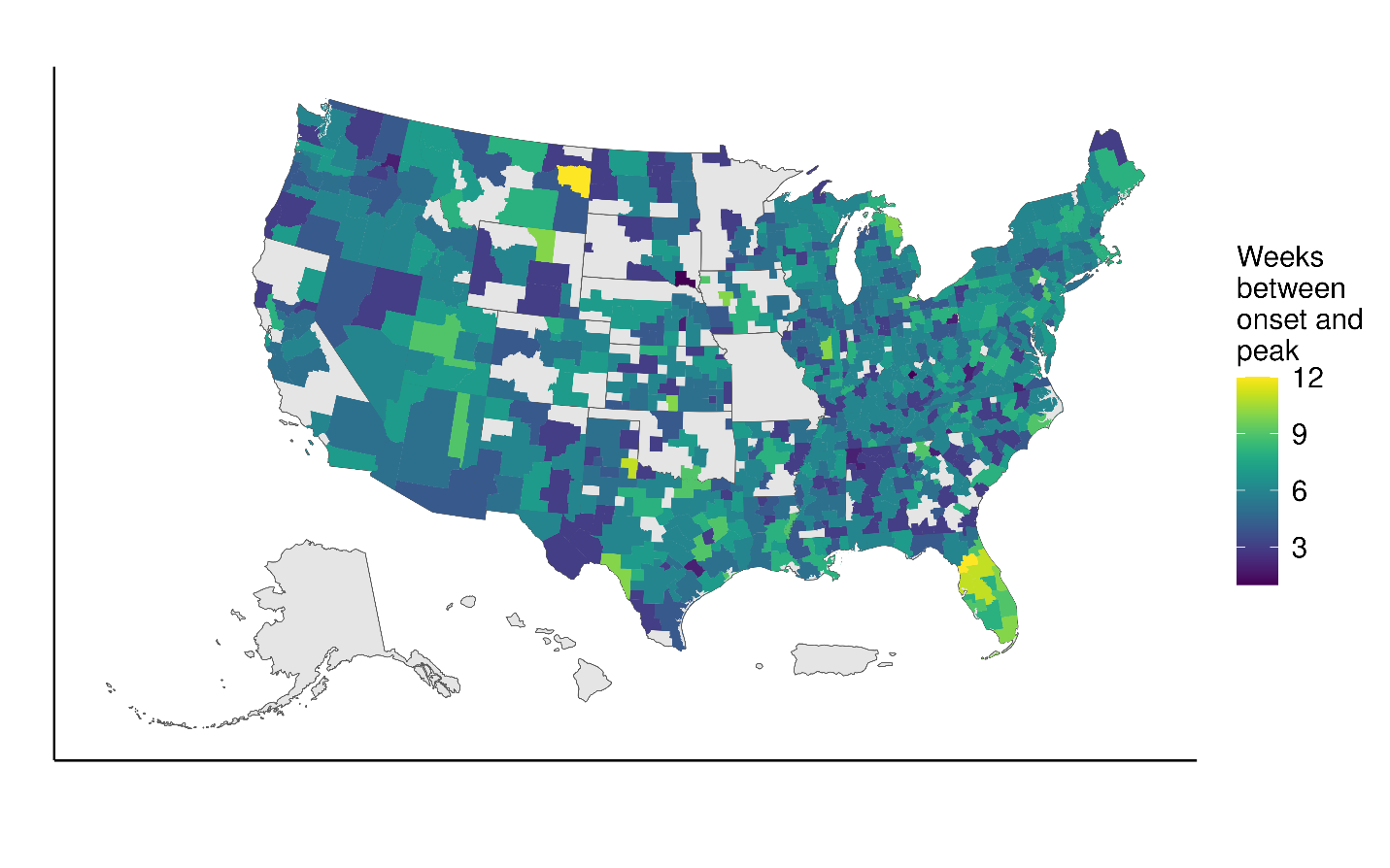

**Figure S4. Estimated time in weeks between estimated onset week and peak week of the 2022/23 influenza season for 724 Health Service Areas (HSAs).** Regions in grey were either absent from the data or excluded during the analysis.

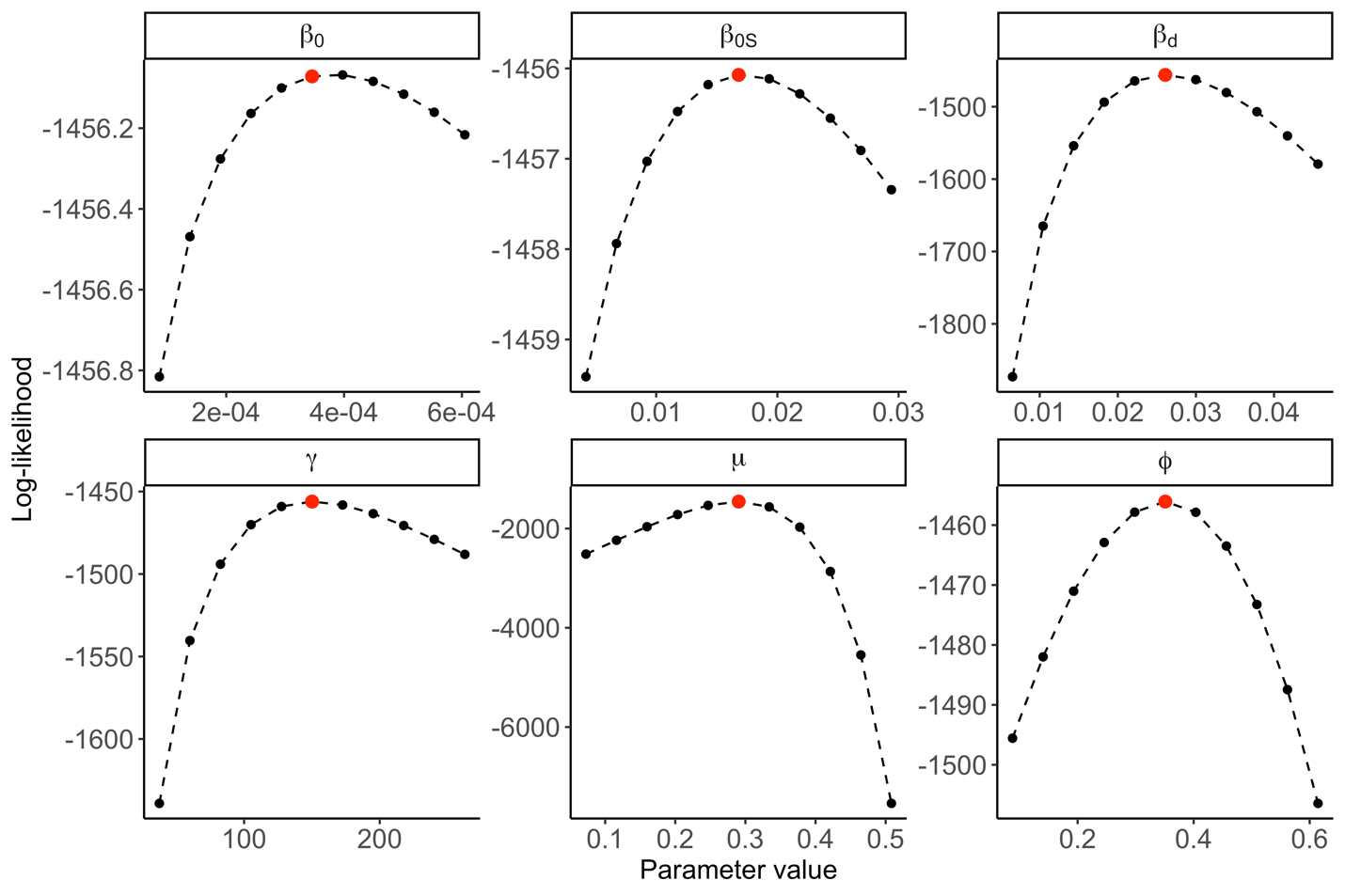
**Figure S6. One-dimensional profile likelihoods for the most parsimonious model with absolute humidity and school opening times.** Red points represent the fitted parameter estimates. Parameters are defined in Table 1.

**References**

[1] tidycensus: Load US Census Boundary and Attribute Data as 'tidyverse' and 'sf'-Ready Data Frames [Internet]. 2025. Available from: <https://walker-data.com/tidycensus/>.

[2] Place and Health - Geospatial Research, Analysis, and Service Programs (GRASP): SVI Data & Documentation Download [Internet]. 2020. Available from: <https://www.atsdr.cdc.gov/place-health/php/svi/svi-data-documentation-download.html>.

[3] Datasets [Internet]. 2025 [cited 14 May 2024]. Available from: <https://cds.climate.copernicus.eu/>.

[4] Parish OO, Putnam TW. Equations for the determination of humidity from dewpoint and psychrometric data. National Aeronautics and Space Administration 1977 [Available from: <https://ntrs.nasa.gov/citations/19770009916>]

[5] Trips by Distance [Internet]. 2024 [cited 14 May 2024]. Available from: <https://data.bts.gov/Research-and-Statistics/Trips-by-Distance/w96p-f2qv/about_data>.

[6] CDC. National Syndromic Surveillance Program (NSSP): About NSSP 2024 [Available from: <https://www.cdc.gov/nssp/php/about/index.html>.
